# Analysis of the genetic diversity of SARS-CoV-2 genomes carrying the Omicron B.1.1.529 mutation

**DOI:** 10.1101/2022.05.21.22275421

**Authors:** Bráulio Wagner Correia da Silva, Pierre Teodosio Felix

## Abstract

In this work, we evaluated the levels of genetic diversity in 95 genomes of the carriers of the Omicron B.1.1.529 mutation in SARS-CoV-2 from South Africa, Asia, Massachusetts-USA, Rhode Island-USA, United Kingdom and Germany. All with 29,996pb extension and recovered from GENBANK and publicly available at the National Center for Biotechnology and Information (NCBI). All gaps and conserved sites were extracted for the construction of a phylogenetic tree and for specific methodologies of estimates of paired F_ST_, Molecular Variance (AMOVA), Genetic Distance, Incompatibility, demographic expansion analyses, molecular diversity and of evolutionary divergence time analyses, always with 20,000 random permutations. The results revealed the presence of only 75 parsimony-informative sites, sites among the 29,996bp analyzed. The analyses based on F_ST_ values, confirmed the absence of distinct genetic structuring with fixation index of 98% and with a greater component of population variation (6%) for a “p” 0.05. Tau variations (related to the ancestry of the groups), did not reveal significant moments of divergence, supported by the incompatible analysis of the observed distribution (τ = 0%). It is safe to say that the large number of existing polymorphisms reflects major changes in the protein products of viral populations in all countries and especially In South Africa. This consideration provides the safety that, because there are large differences between the haplotypes studied, these differences are minimal within the populations analyzed geographically and, therefore, it does not seem safe to extrapolate the results of polymorphism and molecular diversity levels found in the Variant Omicron B.1.529 of SARS-CoV-2 for wild genomes or other mutants. This warns us that, due to their higher transmission speed and infection, possible problems of molecular adjustments in vaccines already in use may be necessary in the near future.

## 1. Introduction

Since the discovery of the *Coronaviridae* family, problems and concerns have been constant in human populations. Initially with the SARS-CoV virus, because of the onset of severe acute respiratory syndrome and which, in a few years later, evolved into the Type SARS-CoV-2, which initiated an endemic disease in the city of Wuhan in China (December 2019), changing its status rapidly to what we recently know as a global pandemic of COVID-19. Today, scientists and researchers reserve greater attention to the emergence of these new forms, called “variants of concern of Sars-CoV-2”, which, as before, have been responsible for new outbreaks, spreading rapidly around the world and causing in addition to serious public health problems, closing borders, trades, schools and a large number of public environments. (Tabibzadeh *et al*, 2021).

Much is speculated about the existence of large differences between wild SARS-CoV-2 and its variants, however, previous studies have shown that, when comparing the primitive form of SARS-CoV-2 with one of its variants (D614G), no increases were observed in the number of deaths, although this variant has a higher rate of dissemination and infection, as well as, no further technical adjustments are made in relation to the vaccines already produced. (Felix *et.al*, 2020a). The low number of deaths caused by the D614G variant may be explained by the low polymorphism found in its genome, such as that detected in the work of the Laboratory of Population Genetics and Computational Evolutionary Biology (LaBECom-UNIVISA) with 38 complete genome sequences of Sars-CoV-2 from South America, where 75 sites were polymorphic in a total of 29.996 bp. (Felix *et.al*, 2020b)

Although studies such as this point to a polymorphism not significant for Sars-CoV-2, the appearance of new variants around the world such as those listed as variants of interest by the CDC (USA) and WHO, such as strains (VOC’s), B.1.1.7 (variant 20I/501Y. V1), P.1 (variant 20J/501Y. V3), B.1.351 (variant 20H/501Y. V2) and B.1.617.2 and (VOI’s), B.1.525, B.1.526, P.2 and B.1.427/B.1.429, with emphasis on attention to the last two variants, has been leaving the world on alert. (Chakraborty *et.al*, 2021). According to a statement by the World Health Organization (WHO), mediated by the International Committee for Virus Taxonomy (ICTV), the total number of cases and deaths from coronavirus justifies concern about the new variants and especially the most recent of them, the variant Ômicron B.1.1.529, discovered on November 26, 2021(Aghamirza *et.al*, 2022), which is coming spreading rapidly and is already confirmed in 194 countries, with effective reproduction numbers that are triple the Delta variant. (Du *et.al*, 2022).

The Ômicron variant specifically makes it difficult to affinity the antibody receptor binding to protein S, resulting in a probable immune and vaccine escape, increasing its transmissibility and likely reinfections by Sars-CoV-2 and, with regard to vaccines, people with incomplete vaccination regimens show a certain degree of protection to this variant, substantially improving their immune response from the application of booster doses. This brings us back to the fact that complete vaccination schemes are the main allies in containing all variants and especially the Omicron variant. (Chekol *et al*, 2022).

Thus, to understand a little more about the molecular diversity of the Omicron variant, 95 sequences of the complete genome of SARS-CoV-2(06 from South Africa, 55 from Asia, 10 from the city of Massachusetts (USA), 02 from the city of Rhode Island (USA), 20 from the United Kingdom and 2 from Germany), were rescued from the National Biotechnology Information Center (NCBI) and submitted to phylum genetic and molecular variance (AMOVA) tests, to understand, in addition to other things, its demographic and spatial expansion.

## 2. Objective

Evaluate possible levels of genetic diversity and polymorphisms existing in 95 mutant haplotypes for the Ômicron B.1.1.529 of SARS-CoV-2.

## 3. Methodology

### 1. Databank

95 sequences of the complete genome of the SARS-CoV-2 virus carrying the Ômicron B.1.1.529 mutation, from South Africa (06), Asia (55), Massachusetts City-USA (10), Rhode Island City-USA (02), United Kingdom (20) and Germany (02), all with 29,996pb extension, were recovered from GENBANK (https://www.ncbi.nlm.nih.gov/labs/gquery/all/?term=omicron&utm_source=Datasets) on April 20, 2022. Once aligned using the MEGA X program (TAMURA *et al*., 2018), ambiguous sites, lost data and gaps, were excluded.

### 2. Genetic Structuring Analyses

Paired F_ST_ estimators, Molecular Variance (AMOVA), Genetic Distance, mismatch, demographic and spatial expansion analyses, molecular diversity and evolutionary divergence time were obtained with the Software Arlequin v. 3.5 (EXCOFFIER *et al*., 2005) using 1000 random permutations (NEI and KUMAR, 2000). The F_ST_ and geographic distance matrices were not compared. All steps of this process are described below:

#### Genetic diversity

Among the routines of LaBECom, this test is used to measure the genetic diversity that is equivalent to the heterozygosity expected in the studied groups. We used for this the standard index of genetic diversity H, described by Nei (1987). Which can also be estimated by the method proposed by PONS and PETIT (1995).

#### Site frequency spectrum (SFS)

According to LaBECom protocols, we used this local frequency spectrum analytical test (SFS), from DNA sequence data that allows us to estimate the demographic parameters of the frequency spectrum. Simulations are made using fastsimcoal2 software, available in http://cmpg.unibe.ch/software/fastsimcoal2/.

#### Molecular diversity indices

Molecular diversity indices are obtained by means of the average number of paired differences, as described by Tajima in 1993, in this test we used sequences that do not fit the model of neutral theory that establishes the existence of a balance between mutation and genetic drift.

#### Calculating theta estimators

Theta population parameters are used in our Laboratory when we want to qualify the genetic diversity of the studied populations. These estimates, classified as Theta Hom – which aim to estimate the expected homozygosity in a population in equilibrium between drift and mutation and the estimates Theta (S) (WATTERSON, 1975), Theta (K) (EWENS, 1972) and Theta (π) (TAJIMA, 1983).

#### Calculation of the distribution of mismatch

In LaBECom, analyses of the mismatch distribution are always performed relating the observed number of differences between haplotype pairs, trying to define or establish a pattern of population demographic behavior, as described already by (ROGERS; HARPENDING, 1992; HUDSON, SLATKIN, 1991; RAY *et al*., 2003, EXCOFFIER, 2004).

#### Pure demographic expansion

This model is always used when we intend to estimate the probability of differences observed between two haplotypes not recombined and randomly chosen, this methodology in our laboratory is used when we assume that the expansion, in a haploid population, reached a momentary balance even having passed through τ generations, of sizes 0 N to 1 N. In this case, the probability of observing the S differences between two non-recombined and randomly chosen haplotypes is given by the probability of observing two haplotypes with S differences in this population (Watterson, 1975).

#### Haplotypic inferences

We use these inferences for haplotypic or genotypic data with unknown gametic phase. Following our protocol, inferences are estimated by observing the relationship between haplotype i and xi times its number of copies, generating an estimated frequency (^pi). With genotypic data with unknown gametic phase, the frequencies of haplotypes are estimated by the maximum likelihood method, and can also be estimated using the expected Maximization (MS) algorithm.

#### Method of Jukes and Singer

This method, when used in LaBECom, allows estimating a corrected percentage of how different two haplotypes are. This correction allows us to assume that there have been several substitutions per site, since the most recent ancestor of the two studied haplotypes. Here, we also assume a correction for identical replacement rates for all four nucleotides A C, G and T.

#### Kimura method with two parameters

Much like the previous test, this fix allows for multiple site substitutions, but takes into account different replacement rates between transitions and transversions.

#### Tamura Method

We at LaBECom understand this method as an extension of the 2-parameter Kimura method, which also allows the estimation of frequencies for different haplotypes. However, transition-transversion relationships as well as general nucleotide frequencies are calculated from the original data.

#### Tajima and Nei method

At this stage, we were also able to produce a corrected percentage of nucleotides for which two haplotypes are different, but this correction is an extension of the Jukes and Cantor method, with the difference of being able to do this from the original data.

#### Tamura and Nei model

As in kimura’s models 2 parameters a distance of Tajima and Nei, this correction allows, inferring different rates of transversions and transitions, besides being able to distinguish transition rates between purines and pyrimidines.

#### Minimum spanning network

To calculate the distance between OTU (operational taxonomic units) from the paired distance matrix of haplotypes, we used a Minimum Spanning Network (MSN) tree, with a slight modification of the algorithm described in Rohlf (1973). We usually use free software written in Pascal called MINSPNET. EXE running in DOS language, previously available at: http://anthropologie.unige.ch/LGB/software/win/min-span-net/.

#### Genotypic data with unknown gametic phase

##### IN algorithm

To estimate haplotypic frequencies we used the maximum likelihood model with an algorithm that maximizes the expected values. The use of this algorithm in LaBECom, allows to obtain the maximum likelihood estimates from multilocal data of gametic phase is unknown (phenotypic data). It is a slightly more complex procedure since it does not allow us to do a simple gene count, since individuals in a population can be heterozygous to more than one locus.

##### ELB algorithm

Very similar to the previous algorithm, ELB attempts to reconstruct the gametic phase (unknown) of multilocal genotypes by adjusting the sizes and locations of neighboring loci to explore some rare recombination.

#### Neutrality tests

##### Ewens-Watterson homozygosis test

We use this test in LaBECom for both haploid and diploid data. This test is used only as a way to summarize the distribution of allelic frequency, without taking into account its biological significance. This test is based on the sampling theory of neutral all links from Ewens (1972) and tested by Watterson (1978). It is now limited to sample sizes of 2,000 genes or less and 1,000 different alleles (haplotypes) or less. It is still used to test the hypothesis of selective neutrality and population balance against natural selection or the presence of some advantageous alleles.

#### Accurate Ewens-Watterson-Slatkin Test

This test created by Slatikin in 1994 and adapted by himself in 1996. is used in our protocols when we want to compare the probabilities of random samples with those of observed samples.

#### Chakraborty’s test of population amalgamation

This test was proposed by Chakrabordy in 1990, serves to calculate the observed probability of a randomly neutral sample with a number of alleles equal to or greater than that observed, it is based on the infinite allele model and sampling theory for neutral Alleles of Ewens (1972).

#### Tajima Selective Neutrality Test

We use this test in our Laboratory when DNA sequences or haplotypes produced by RFLP are short. It is based on the 1989 Tajima test, using the model of infinite sites without recombination. It commutes two estimators using the theta mutation as a parameter.

#### *FS FU* Test of Selective Neutrality

Also based on the model of infinite sites without recombination, the FU test is suitable for short DNA sequences or haplotypes produced by RFLP. However, in this case, it assesses the observed probability of a randomly neutral sample with a number of alleles equal to or less than the observed value. In this case the estimator used is θ.

#### Methods that measure interpopulation diversity

##### Genetic structure of the population inferred by molecular variance analysis (AMOVA)

This stage is the most used in the LaBECom protocols because it allows to know the genetic structure of population measuring their variances, this methodology, first defined by Cockerham in 1969 and 1973) and, later adapted by other researchers, is essentially similar to other approaches based on analyses of gene variance variance, but takes into account the number of mutations haplotypes. When the population group is defined, we can define a particular genetic structure that will be tested, that is, we can create a hierarchical analysis of variance by dividing the total variance into covariance components by being able to measure intra-individual differences, interindividual differences and/or interpopulation allocated differences.

#### Minimum Spanning Network (MSN) among haplotypes

In LaBECom, this tree is generated using the operational taxonomic units (OTU). This tree is calculated from the matrix of paired distances using a modification of the algorithm described in Rohlf (1973).

#### Locus-by-locus AMOVA

We performed this analysis for each locus separately as it is performed at the haplotypic level and the variance components and f statistics are estimated for each locus separately generating in a more global panorama.

#### Paired genetic distances between populations

This is the most present analysis in the work of LaBECom. These generate paired F_ST_ parameters that are always used, extremely reliably, to estimate the short-term genetic distances between the populations studied, in this model a slight algorithmic adaptation is applied to linearize the genetic distance with the time of population divergence (Reynolds *et al*. 1983; Slatkin, 1995).

#### Reynolds Distance (Reynolds *et al*. 1983)

Here we measured how much pairs of fixed N-size haplotypes diverged over t generations, based on F_ST_ indices.

#### Slatkin’s linearized F_ST_’s (Slatkin 1995)

We used this test in LaBECom when we want to know how much two Haploid populations of N size diverged t generations behind a population of identical size and managed to remain isolated and without migration. This is a demographic model and applies very well to the phylogeography work of our Laboratory.

#### Nei’s average number of differences between populations

In this test we assumed that the relationship between the gross (D) and liquid (AD) number of Nei differences between populations is the increase in genetic distance between populations (Nei and Li, 1979).

#### Relative population sizes: divergence between populations of unequal sizes

We used this method in LaBECom when we want to estimate the time of divergence between populations of equal sizes (Gaggiotti and Excoffier, 2000), assuming that two populations diverged from an ancestral population of N0 size a few t generations in the past, and that they have remained isolated from each other ever since. In this method we assume that even though the sizes of the two child populations are different, the sum of them will always correspond to the size of the ancestral population. The procedure is based on the comparison of intra and inter populational (π’s) divers that have a large variance, which means that for short divergence times, the average diversity found within the population may be higher than that observed among populations. These calculations should therefore be made if the assumptions of a pure fission model are met and if the divergence time is relatively old. The results of this simulation show that this procedure leads to better results than other methods that do not take into account unequal population sizes, especially when the relative sizes of the daughter populations are in fact.

#### Accurate differentiation population tests

We at LaBECom understand that this test is an analog of fisher’s exact test in a 2×2 contingency table extended to a rxk contingency table. It has been described in Raymond and Rousset (1995) and tests the hypothesis of a random distribution of k different haplotypes or genotypes among r populations.

#### Assignment of individual genotypes to populations

Inspired by what had been described in Paetkau *et al* (1995, 1997) and Waser and Strobeck (1998) this method determines the origin of specific individuals, knowing a list of potential source populations and uses the allelic frequencies estimated in each sample from their original constitution.

#### Detection of loci under selection from F-statistics

We use this test when we suspect that natural selection affects genetic diversity among populations. This method was adapted by Cavalli-Sforza in 1966 from a 1973 work by Lewontin and Krakauer.

## 4. Results

All 95 sequences of the Mutant SARS-CoV-2 Ômicron B.1.1.529 virus from South Africa, Asia, Massachusetts-USA, Rhode Island-USA, United Kingdom and Germany, revealed the presence of only 75 polymorphic and informative parsimony sites among the 29,996bp analyzed (Table 1).

**Table 1.** Description of the percentage of conservation in all groups of sequences analyzed in this study.

| Geographic location | Number of haplotypes | Polymorphism level | Conservation percentage |
| --- | --- | --- | --- |
| SOUTH AFRICA OMICRON | 06 haplotypes | Highly polymorphic | 8,25% |
| ÓMICRON ASIA | 55 haplotypes | Highly polymorphic | 0,05% |
| MASSACHUSETTS-US OMICRON | 10 haplotypes | Highly polymorphic | 0,32% |
| RHODE ISLAND-USA OMICRON | 02 haplotypes | Highly polymorphic | 0,18% |
| OMICRON UNITED KINGDOM | 20 haplotypes | Highly polymorphic | 0,22% |
| ÓMICRON SCHLESWIG HOLSTEIN-GERMANY | 02 haplotypes | Highly polymorphic | 0,12% |

Using the UPGMA method, for the 75 parsimony-informative sites, it was possible to understand that the 34 haplotypes from Europe and American continents comprised 3 distinct groups, and it is even possible that there are haplotype sharing between the studied countries (Figure 2).

**Figure 2.**
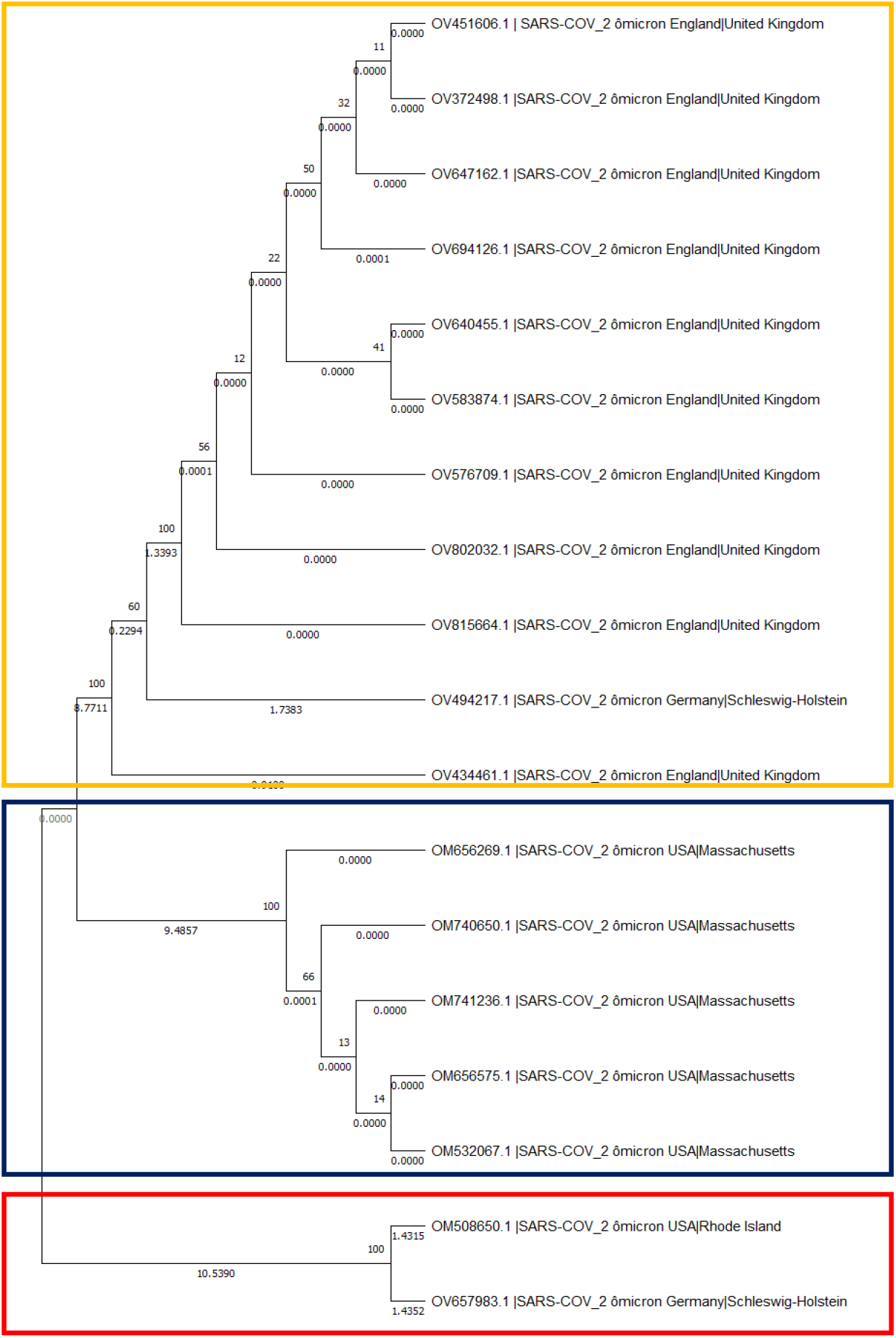
Evolutionary analysis by the maximum likelihood method. The evolutionary history was inferred using the maximum likelihood method and the 3-parameter Tamura model[1]. The tree with the highest probability of logging (−1366.35) is shown. The percentage of trees in which the associated dollar sums group together is shown next to the branches. The initial trees for heuristic research were obtained automatically by applying the Join-Join and BioNJ algorithms to an array of distances in estimated pairs using the Tamura 3 parameter model and then selecting the topology with a higher log probability value. This analysis involved 38 nucleotide sequences. The evolutionary analyses were performed in MEGA X.

### Molecular Variance Analysis (AMOVA) and Genetic Distance

Analyses based on F_ST_ values confirmed the presence of several distinct genetic entities with a fixation index of 98% and with a larger component of population variation of 2% to a p lower than 0.05. Significant evolutionary divergences were observed within and between groups (Table 2) and little genetic similarity between the groups that make up the American and European countries, as well as a greater evolutionary divergence between the sequences that make up the United Kingdom Group (Table 3, Table 4, Figure 3 and Figure 4).

**Table 2.** Components of haplotypic variation and paired F_ST_ value.

| **Table 2.** Components of haplotypic variation and paired $F_{ST}$ value. | | | |
| --- | --- | --- | --- |
| **AMOVA design and results (average polymorphic loci only):** |  |  |  |
| **Source of Variation** | **Sum of squares** | **Variance components** | **Percentage variation** |
| **Among Populations** | 291.756 | 5.93488 | 97.97883 |
| **Within Populations** | 5.400 | 0.12243 | 2.02117 |
| **Total** | 297.156 | 6.05731 |  |
| Average F-Statistics over all loci Fixation Indices ($F_{ST}$) : **0.97979** | | | |

**Table 3.**
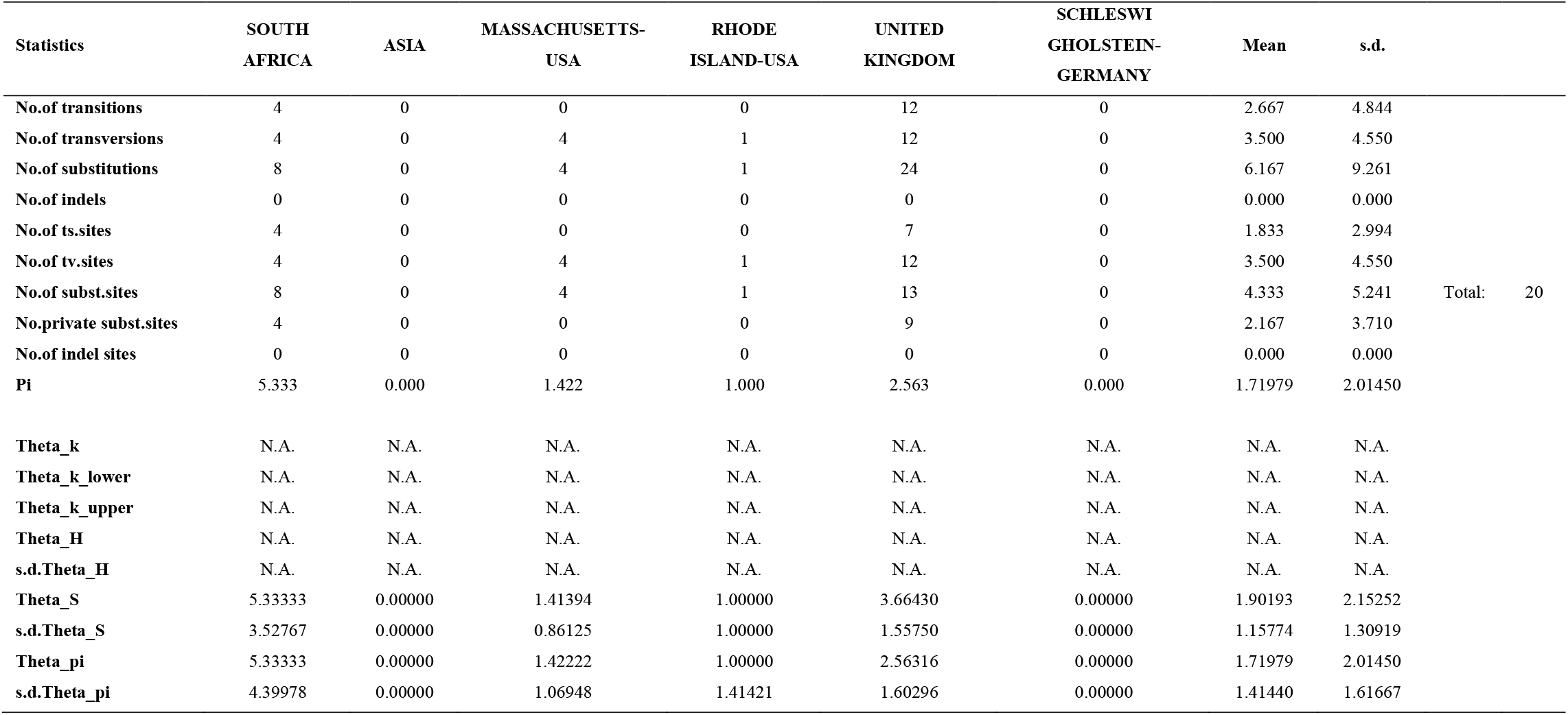
Molecular Diversity Indices of the parsimony-informative sites of Mutant SARS-CoV-2 Ômicron B.1.1.529 virus

**Table 4.** Neutrality Tests for the sequences of Mutant SARS-CoV-2 Ômicron B.1.1.529 virus

| Statistics | SOUTH AFRICA | ASIA | MASSACHUSETTS-USA | RHODE ISLAND-USA | UNITED KINGDOM | SCHLESWI GHOLSTEIN-GERMANY | Mean | s.d. |
| --- | --- | --- | --- | --- | --- | --- | --- | --- |
| **Ewens-Watterson test** |  |  |  |  |  |  |  |  |
| Sample size | 3 | 56 | 10 | 2 | 20 | 2 | 15.50000 | 21.03093 |
| No.of alleles(unchecked) | 3 | 56 | 10 | 2 | 20 | 2 | 15.50000 | 21.03093 |
| Observed F value | N.A | N.A | N.A | N.A. | N.A | N.A. | N.A | N.A. |
| Expected F value | N.A | N.A | N.A | N.A | N.A | N.A | N.A | N.A. |
| Watterson test: PR (Rand<br>E<=obs F) | N.A | N.A | N.A | N.A | N.A. | N.A. | N.A | N.A. |
| Slatkin's exact test P-value | N.A | N.A. | NA | N.A. | N.A. | N.A | N.A | N.A. |
| **Chakraborty's test** |  |  |  |  |  |  |  |  |
| Sample size | 3 | 56 | 10 | 2 | 20 | 2 | 15.50000 | 21.03093 |
| No.of alleles (unchecked) | 3 | 56 | 10 | 2 | 20 | 2 | 15.50000 | 21.03093 |
| Obs.homozygosity | 0.000000 | 0.0000 | .000000 | .000000 | .000000 | .000000 | .000000. | 00000 |
| Exp.no.ofalleles | 2.569381 | 0.0003 | .455981 | .500006 | .049901 | .000002. | 595881. | 94758 |
| P(k or more alleles) | N.A | N.A. | N.A | N.A | N.A. | N.A | N.A. | N.A. |
| **Tajima's D test** |  |  |  |  |  |  |  |  |
| Sample size | 3 | 56 | 10 | 2 | 20 | 2 | 15.50000 | 21.03093 |
| S | 8 | 0 | 4 | 1 | 13 | 0 | 4.33333 | 5.24087 |
| Pi | 5.33333 | 0.00000 | 1.42222 | 1.00000 | 2.56316 | 0.00000 | 1.71979 | 2.01450 |
| Tajima's D | 0.00000 | 0.00000 | 0.02248 | 0.00000- | 1.09005 | 0.00000 | -0.17793 | 0.44694 |
| Tajima's D p-value | 0.90000 | 1.00000 | 0.60000 | 1.00000 | 0.20000 | 1.00000 | 0.78333 | 0.32506 |
| **Fu's FS test** |  |  |  |  |  |  |  |  |
| No.of alleles (unchecked) | 3 | 56 | 10 | 2 | 20 | 2 | 15.50000 | 21.03093 |
| Theta_pi | 5.33333 | 0.00000 | 1.42222 | 1.00000 | 2.56316 | 0.00000 | 1.71979 | 2.01450 |
| Exp.no.of alleles | 2.56938 | 1.00000 | 3.45598 | 1.50000 | 6.04990 | 1.00000 | 2.59588 | 1.94758 |
| FS | 0.45758 | 0.00000 | -12.7042 | -26.4448 | 0.00000 | 0.00000 | 0.00000 | 0.00000 |
| FS p-value | 0.50000 | 1.00000 | 0.00000 | 0.40000 | 0.00000 | 1.00000 | 0.48333 | 0.44907 |

**Figure 3.**
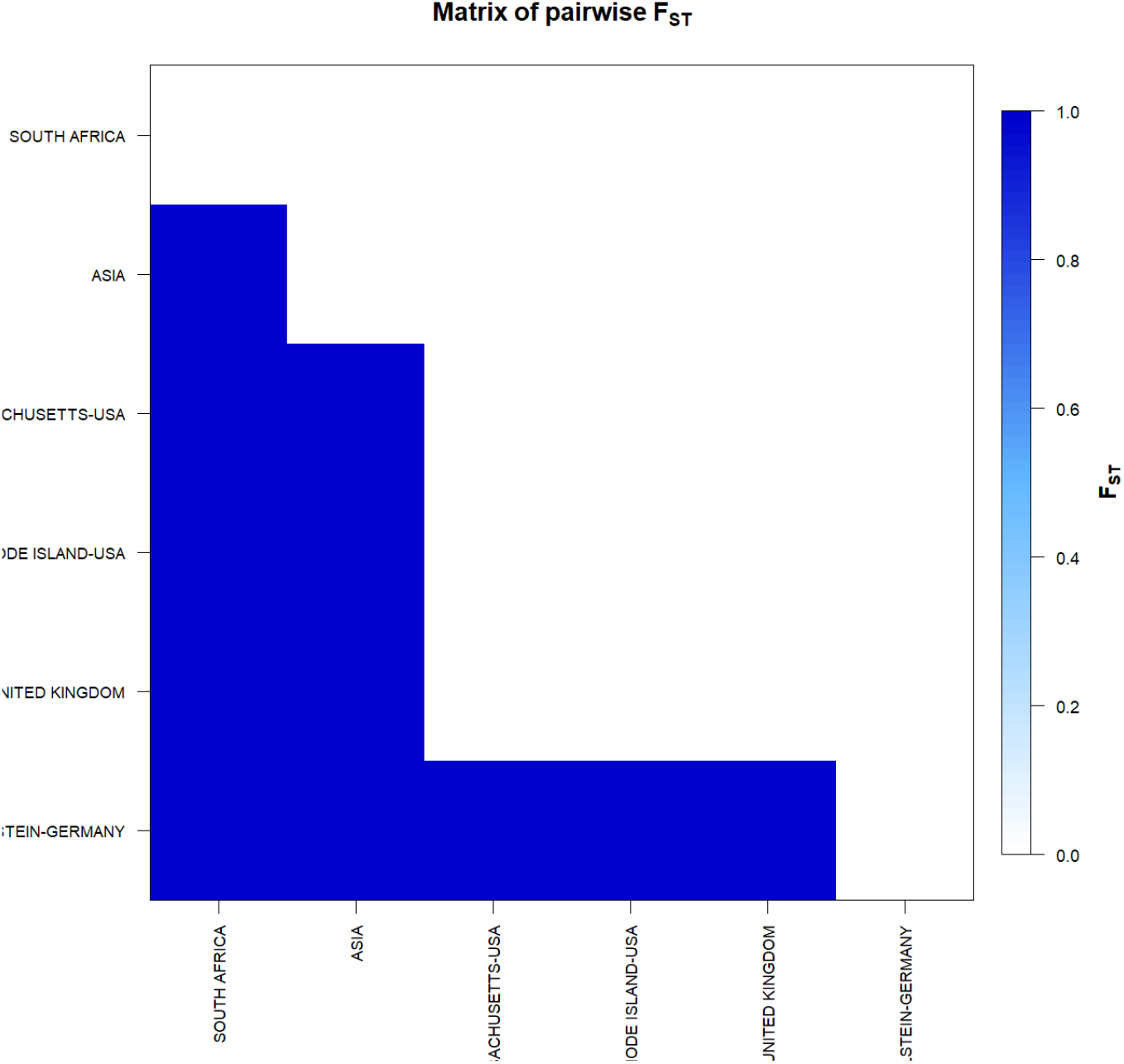
F_ST_-based genetic distance matrix between for the sequences of Mutant SARS-CoV-2 Ômicron B.1.1.529 virus. * Generated by the statistical package in R language using the output data of the Software Arlequin version 3.5.1.2

**Figure 4.**
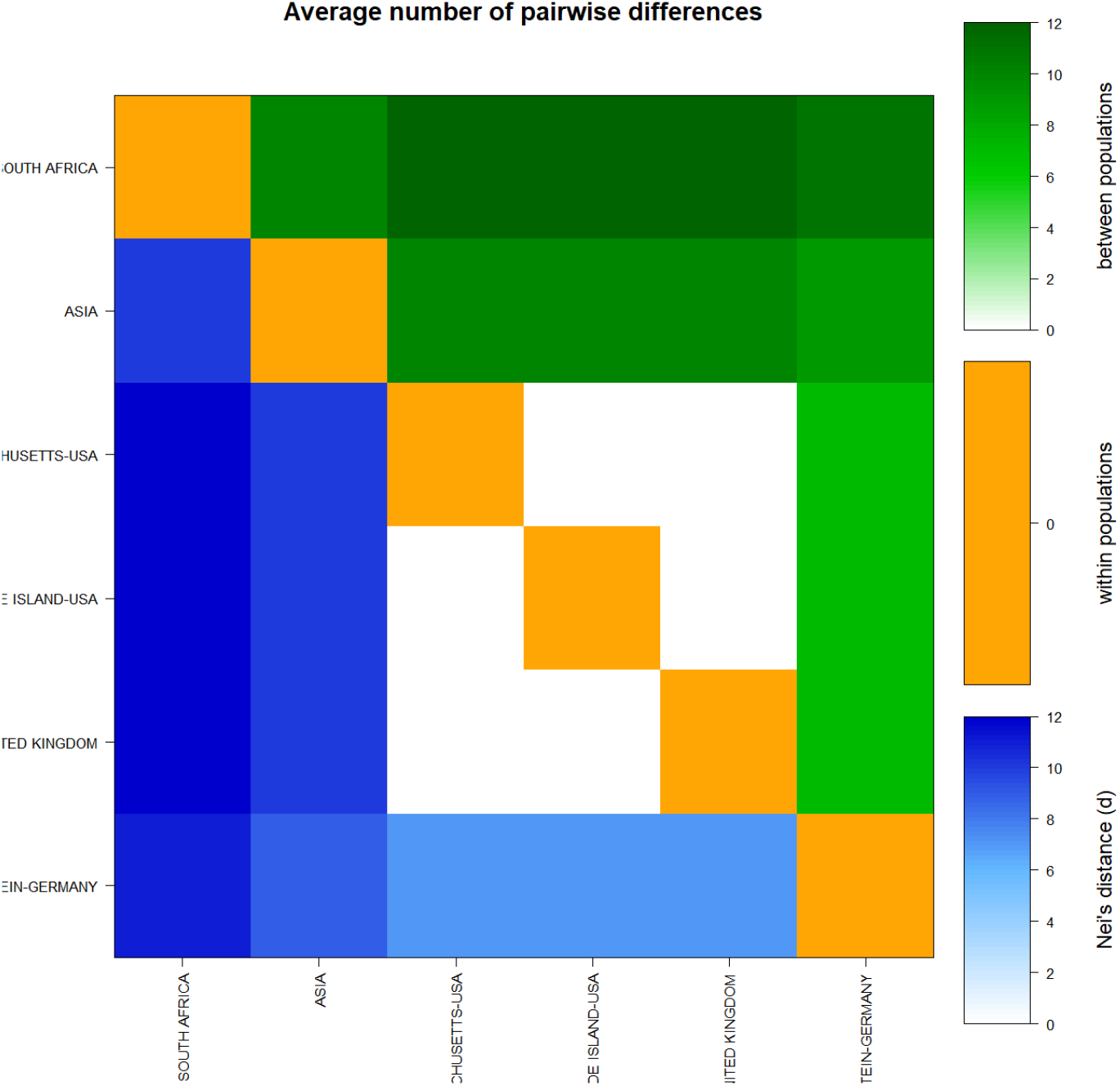
Matrix of paired differences between the populations studied: between the groups; within the groups; and Nei distance for the sequences of Mutant SARS-CoV-2 Ômicron B.1.1.529 virus

*Tau* variations (related to the ancestry of all groups) revealed significant moment of divergence, supported by incompatible analysis of the observed distribution (τ = 42%) and with constant mutation rates between localities.

Molecular analyses estimated by φ reflected a significant level of mutations among all haplotypes (transitions and transversions). Indel mutations (insertions or additions) were found more frequently in the UK group (Table 3). The Tajima and FS de Fu tests showed disagreements between the estimates of general φ and π, but with positive and weakly significant values, indicating, once again, the presence of population expansions for all groups (Table 4). The irregularity index (R= Raggedness) with parametric bootstrap simulated new values φ for before and after a supposed demographic expansion and, in this case, assumed a value greater than zero for all groups, with the exception of the population of South Africa(Table 5), probably due to the small number of haplotypes analyzed (Figure 6 and 7).

**Table 5.**
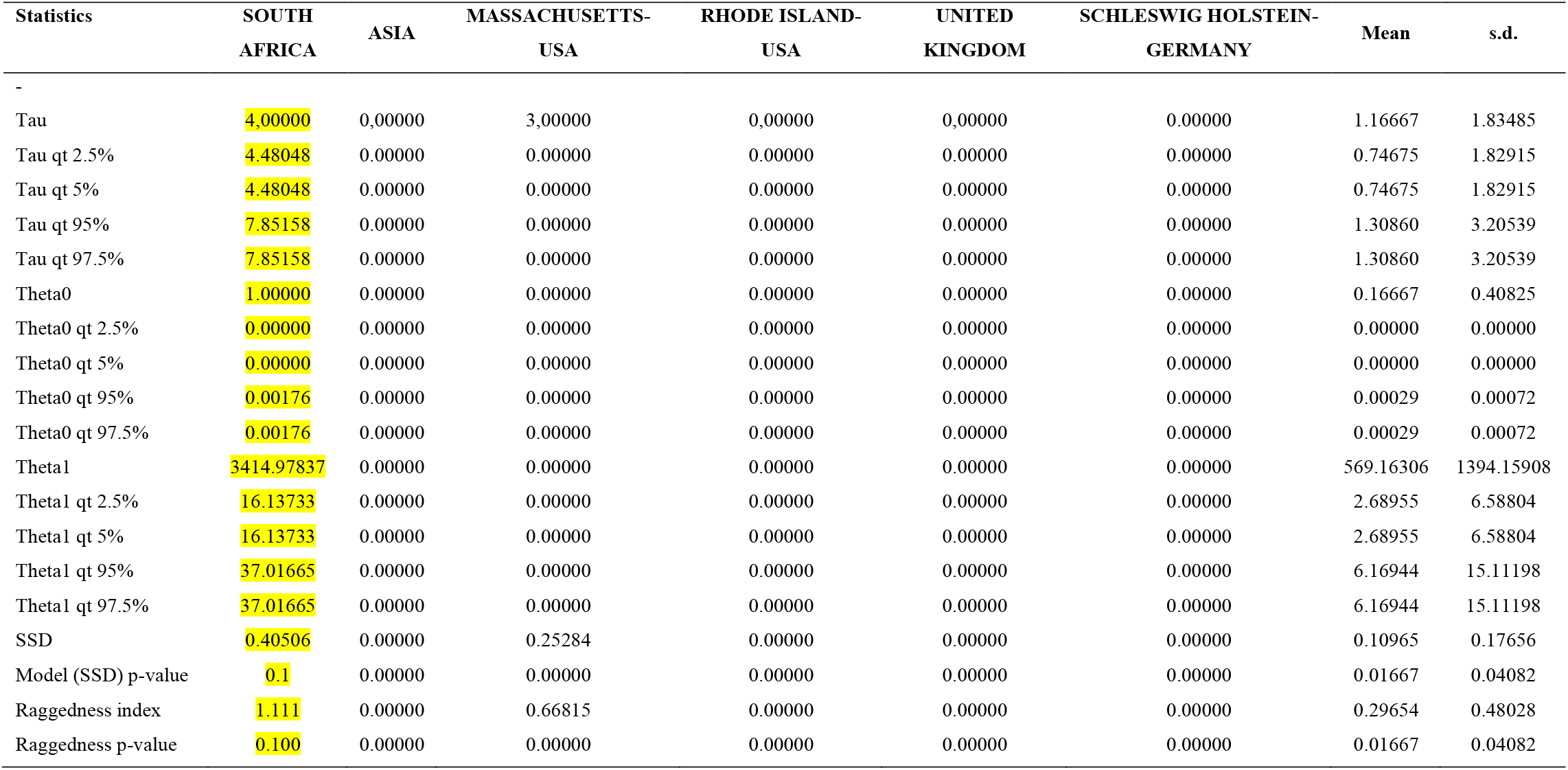
Demographic expansion simulations based on the τ, θ, and M indices for the sequences of Mutant SARS-CoV-2 Ômicron B.1.1.529 virus

A significant similarity was also evidenced for the time of genetic evolutionary divergence among all populations; supported by variations, incompatible analyses and demographic expansion analyses (Figure 5).

**Figure 5.**
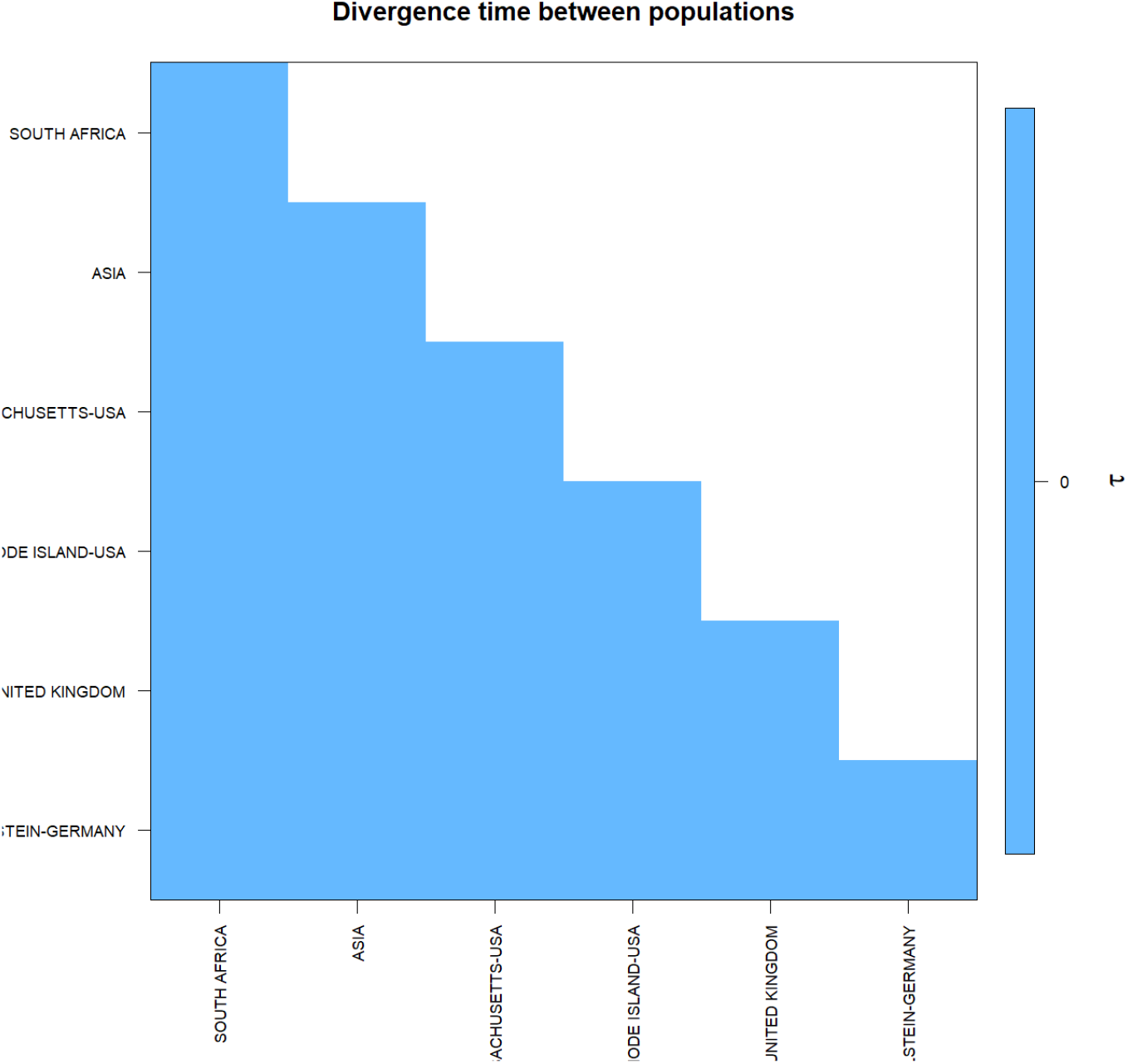
Matrix of divergence time between the populations studied: between the groups; within the groups; and Nei distance for the sequences of Mutant SARS-CoV-2 Ômicron B.1.1.529 virus.

**Figure 6.**
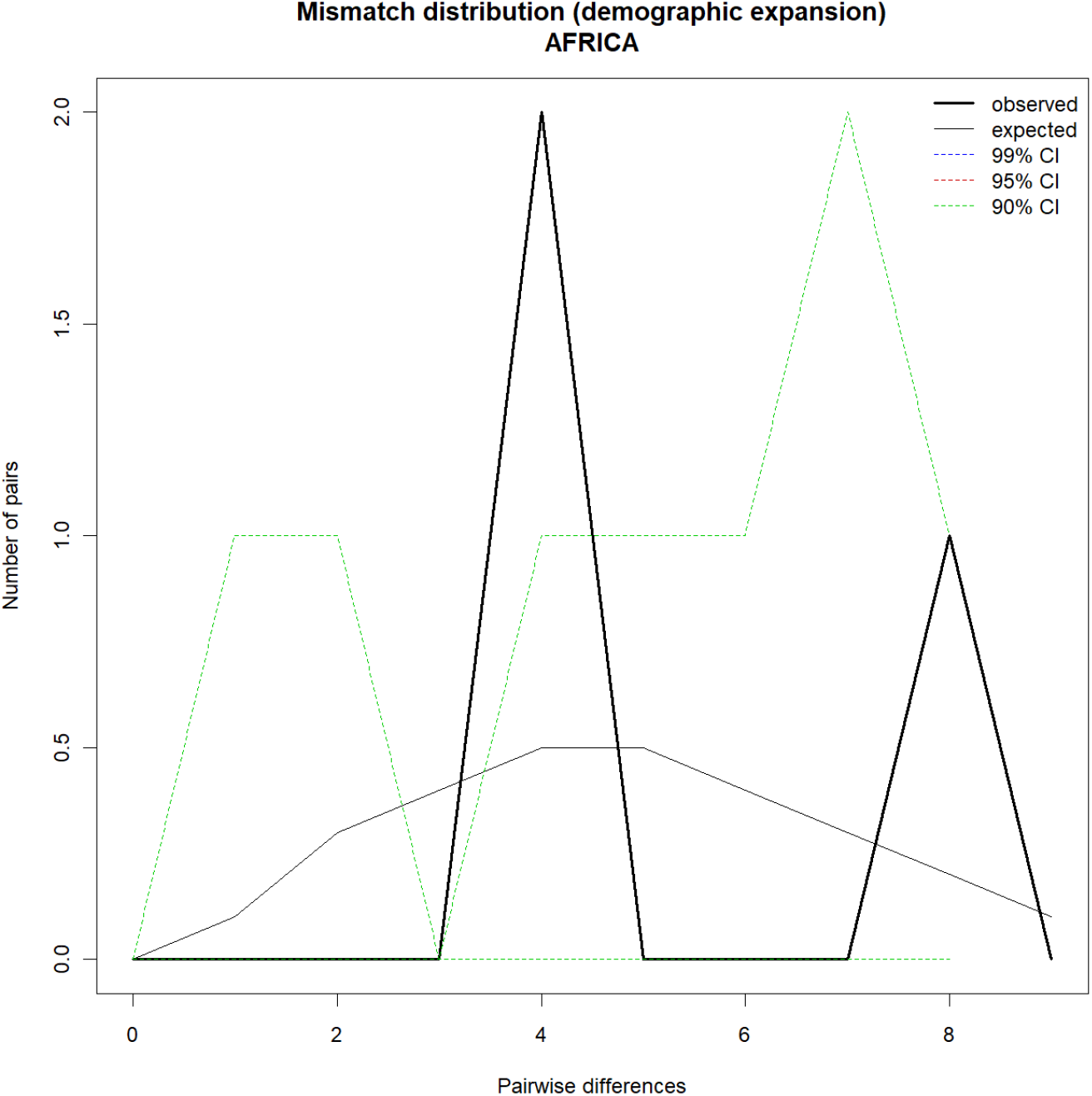
Demographic Expansion for SARS-CoV-2 carrier sequences of Mutant SARS-CoV-2Ômicron B.1.1.529 in South Africa. *Graphs Generated by the R-language statistical package using the output data from version 3.5.1.2 of the Arlequin Software.

**Figure 7.**
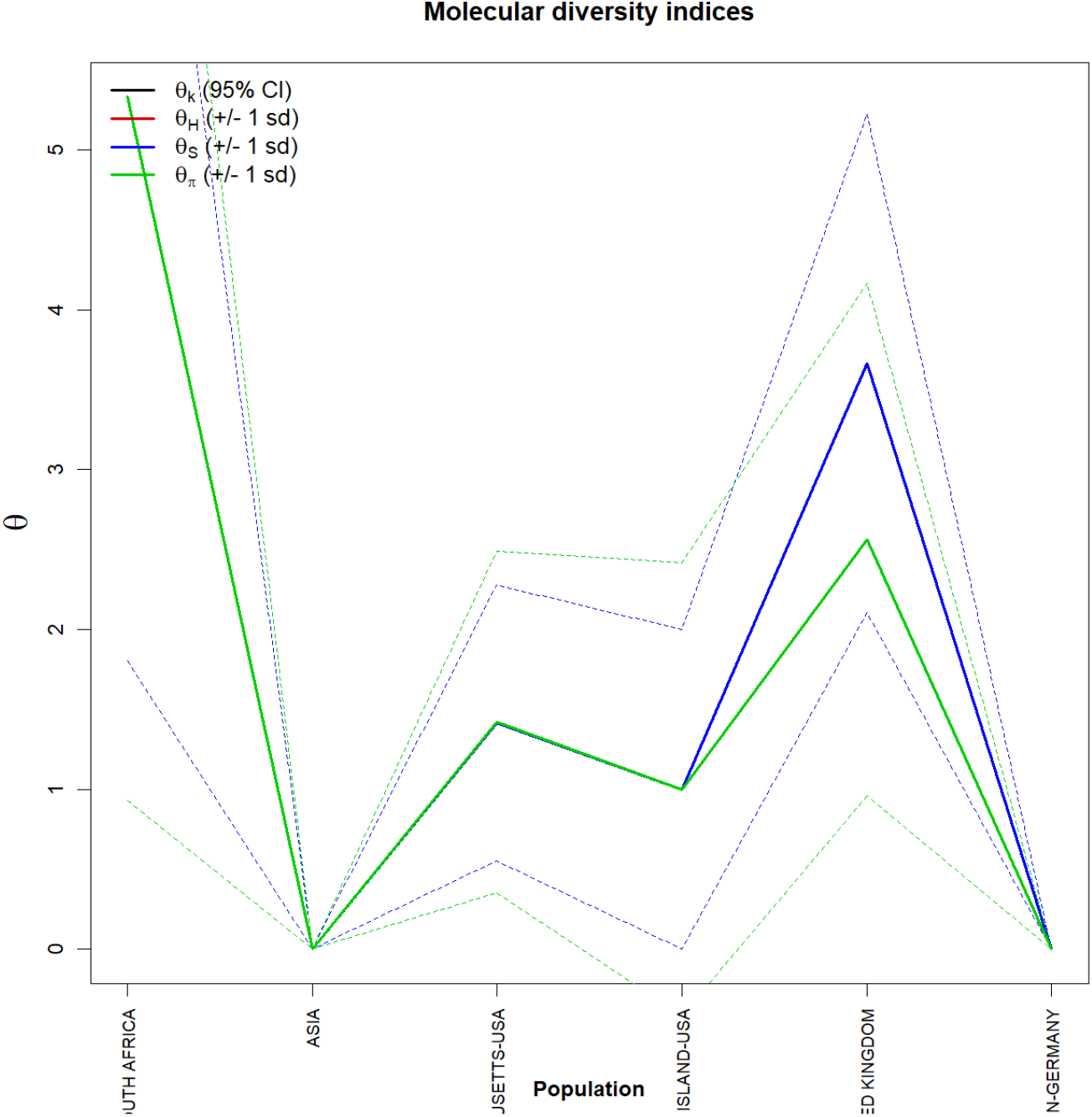
Graph of molecular diversity indices for SARS-CoV-2 carrier sequences of the Mutant SARS-CoV-2 Ômicron B.1.1.529. In the graph the values φ: (φk) Relationship between the expected number of alllos (k) and the sample size; (φH) Expected homozygosity in a balanced relationship between drift and mutation; (φS) Relationship between the number of segregating sites (S), sample size (n) and non-recombinant sites; (φπ) Relationship between the average number of paired differences (π) and φ. * Generated by the statistical package in R language using the output data of the Software Arlequin version 3.5.1.2.

**Figure 8.**
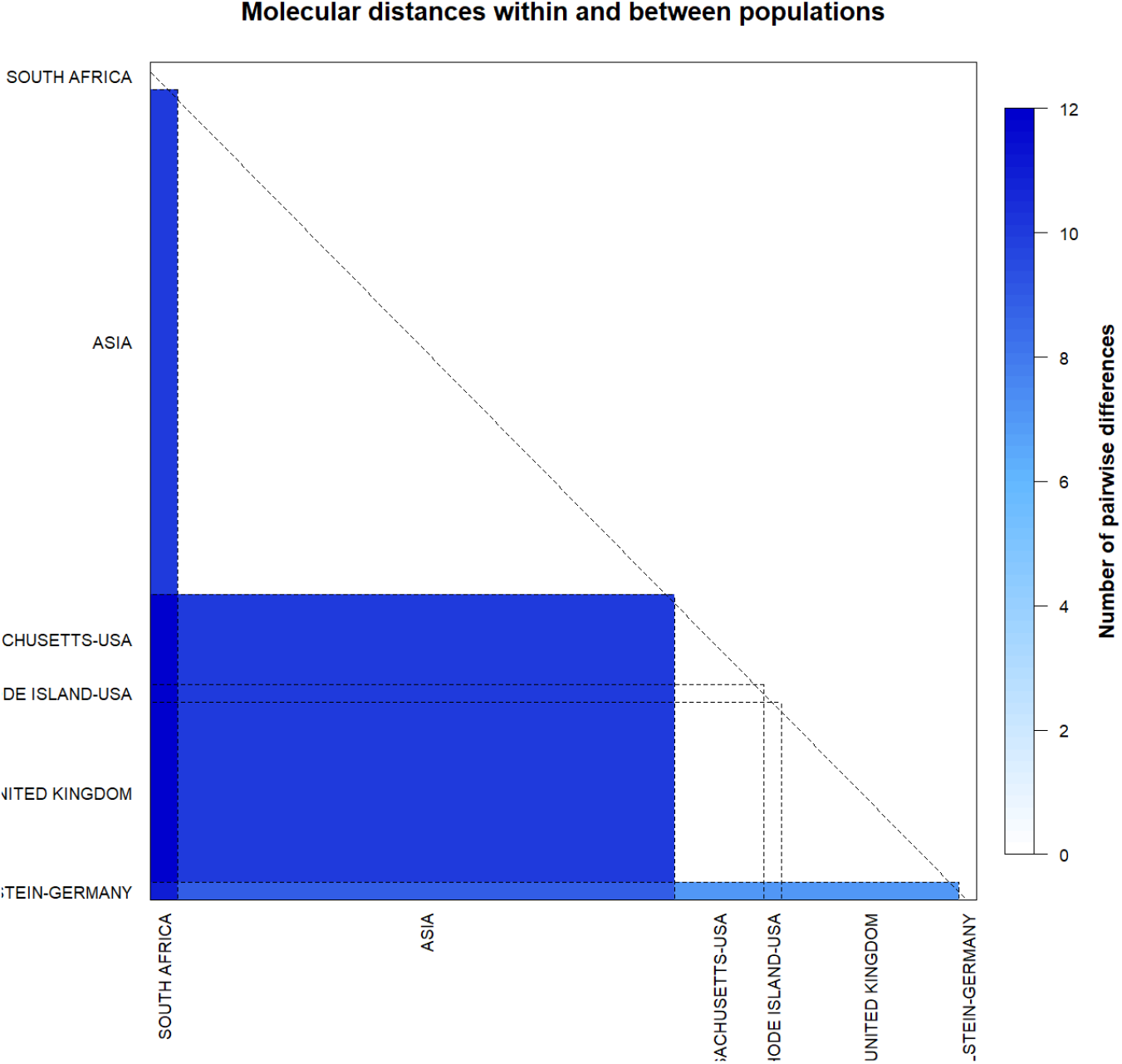
Matrix of Molecular Distances within and between haplotypes of the Mutant SARS-CoV-2 Ômicron B.1.1.529. *Generated by the R-language statistical package using the output data from the Arlequin version 3.5.1.2 software.

## 5. Discussion

As the use of genetic diversity study methodologies not yet used in this customized PopSet for genomes of the Variant Ômicron B.1.1.529 of SARS-CoV-2, it was possible to detect the immense genetic diversity among all haplotypes studied, not evienciarelationship between all localities. The few parsimony-informative sites of all viral genomes contain few inter-haplotypic variations with a few ecessoes such as the UK finger and Massasuchets populations. Perhaps, with the insertion of more haplotypes for all regions studied in the databases, this configuration may change. The groups described here presented null patterns of genetic structuring diverging from the results obtained by Felix *et al*, 2020 for wild populations of SARS-CoV-2 for Latin American countries.

These data suggest that the lack of structure present in the tavez amaostras do not directly relate to the genetic flow. Cases like this have also been supported by simple phylogenetic pairing methodologies, such as UPGMA, which in this case, with the lack of a discontinuous pattern of genetic divergence between the groups, does not support the idea of possible isolations resulting from past fragmentation events, especially when we observe a not-so-numerous amount of branches in the tree generated and with very few mutational steps.

These few mutations have not yet been corrected by drift or the founding effect, which does not accompany the behavior of dispersion and/or loss of intermediate haplotypes over generations. The values found for the genetic distance support the absence of this continuous pattern and low divergence between the groups studied, since they considered important the minimum differences between the groups, when the hapltypes between them were exchanged, as well as the inference of values greater than or equal to those observed in the proportion of these permutations, including the p-value of the test.

The discrimination of the 95 genetic entities was also perceived by almost null and not hierarchical inter haplotypic variations in all components of covariance: by their intra- and inter-individual differences or by their intra- and intergroup differences, generating a dendrogram that supports the idea that the significant similarities found among all populations studied, for example, they have not yet been shared either in their form or in their number, since the result of estimates of the mean evolutionary divergence found within the groups are high..

Based on the low level of haplotypic sharing, tests that measure the relationship between genetic distance and geographic distance, such as the Mantel test were dispensed with. The estimator φ, although extremely sensitive to any form of molecular variation (FU, 1997), did not support the uniformity between the results found by all the methodologies employed, and can be interpreted as a lack of phylogenetic confirmation for a consensus in the conservation of genomes of the variant Ômicron B.1.1.529 of SARS-CoV-2 and, therefore, it is safe to affirm that the large number of existing polymorphisms reflects in large changes in protein products of viral populations in all the locations studied. This consideration provides the safety that, because there are large differences between the haplotypes studied, these differences are minimal within the populations analyzed geographically and, therefore, it does not seem safe to extrapolate the results of polymorphism and molecular diversity levels found in the Variant Omicron B.1.1.529 of SARS-CoV-2 for wild genomes or other mutants, even supporting the discussion of the existence of great differences in the protein products of this mutant in relation to the others. This warns us that, due to their higher transmission speed and infection, possible problems of molecular adjustments in vaccines already in use may be necessary in the near future.

## Data Availability

All data produced in the present study are available upon reasonable request to the authors

